# Clinical practice guidelines to prevent hospital falls: protocol for a systematic review

**DOI:** 10.1101/2023.07.27.23293232

**Authors:** Jonathan P. McKercher, Casey L. Peiris, Anne-Marie Hill, Meg E. Morris

## Abstract

**Background:** Hospital falls are a major global problem responsible for considerable morbidity, mortality, and healthcare costs. Clinical practice guidelines for hospital falls prevention, facilitate the implementation of falls prevention research into practice, through the provision of evidence-based recommendations to mitigate the risk of falls. The review aims to identify, synthesise, appraise, and compare published clinical practice guidelines on hospital falls prevention.

**Methods and analysis:** The databases of MEDLINE, Embase, CINAHL, Cochrane CENTRAL, PEDro, Web of Science, Infobase of Clinical Practice Guidelines, and Epistemonikos will be searched for clinical practice guidelines published between 1993 and 2023. The quality of the guidelines will be evaluated using the Appraisal of Guidelines for Research and Evaluation Global Rating Scale and Recommendation EXcellence tools. A thematic synthesis will be performed on the content of the included clinical practice guidelines. The synthesis findings will be graded using the Grading of Recommendations Assessment, Development and Evaluation Confidence in Evidence from Reviews of Qualitative Research approach.

**Ethics and dissemination:** Ethics approval is not required for the review. The review will be submitted for publication to a peer-reviewed journal.

## BACKGROUND

Hospital falls and their associated injuries are a major and persistent problem across the globe.^1 2^ Hospital falls are responsible for significant healthcare costs and preventable morbidity and mortality.^2-4^ Clinical practice guidelines are used to implement evidence from research into clinical practice in order to improve patient outcomes.^5^ Clinical practice guidelines on hospital falls prevention have a focus on mitigating the risk of falls in hospitals through implementing evidence-based, best-practice recommendations.^5^ The systematic review aims to identify, synthesise, appraise, and compare published clinical practice guidelines on hospital falls prevention.

## METHODS AND ANALYSIS

This protocol has been guided by the Preferred Reporting Items for Systematic Reviews and Meta-Analysis Protocols (PRISMA-P) recommendations.^6^ The systematic review will be registered on the international prospective register of systematic reviews (PROSPERO),^7^ and will be written in compliance with the Preferred Reporting Items for Systematic Reviews and Meta-Analyses (PRISMA) recommendations.^8^ Data synthesis will be guided by the Enhancing Transparency in Reporting the Synthesis of Qualitative Research (ENTREQ).^9^

### Eligibility criteria

Clinical guidelines will be included in the review if they: (i) are published clinical practice guidelines; (ii) pertain to falls prevention (including falls screening, comprehensive assessment of falls or falls related injuries or interventions; and (iii) pertain to adults or older adults in hospital. Clinical practice guidelines will be included if they originate from countries included in The United Nations regional group of Western European and Other Groups (WEOG).^10 11^ This group of countries, which includes the United States of America, Canada, United Kingdom, Australia and others, was selected as they have comparable healthcare systems.

Guidelines will be excluded if they are limited to infants, children, or adolescent falls prevention; contain recommendations restricted to outpatient, residential aged-care, community, or home settings or if the mode of delivery is solely online. The following will be excluded: guidelines pertaining only to occupational or sport related falls, systematic reviews, policies, guidelines where the full text is unavailable, previous versions of included guidelines (i.e., not the most up-to-date version), and guidelines that are older than 30 years. A 30-year date limit was set to reflect one of the earliest formally endorsed clinical practice guidelines for managing falls.^12^

### Identification and selection of included guidelines

Search terms and synonyms were developed for the following key concepts: clinical practice guidelines, falls prevention, and hospitals. The search strategy was reviewed by a senior research librarian. The databases will include MEDLINE, Embase, CINAHL, Cochrane CENTRAL, PEDro, Web of Science, Infobase of Clinical Practice Guidelines, and Epistemonikos. These databases will be searched from 1 January 1993. An example search is included in the Supplemental Material. A grey literature search will also be performed for organisations including the National Health and Medical Research Council (NHMRC) Clinical Practice Guidelines, Guidelines International Network, Australian Commission on Safety and Quality in Health Care (ACSQHC), National Institute for Health and Care Excellence (NICE), Centers for Disease Control and Prevention (CDC) and the National Health Service (NHS). The database and grey literature searches will be supplemented by reviewing the reference lists of systematic reviews of clinical practice guidelines on falls prevention and the reference lists of the included guidelines. An email will be sent to researchers specialising in hospital falls prevention to find any guidelines not retrieved during the database, grey literature, or supplemental searches.

The search results will be uploaded to Covidence, a review management software,^13^ for screening. The duplicates will first be deleted before the titles and abstracts will be screened by two reviewers. The full texts of those not excluded will then be screened by two reviewers. Reasons for exclusion will be recorded. The two reviewers will meet to reach consensus on the discrepancies that arise during each screening stage. A PRISMA compliant flow diagram will be used to display the selection process of the included guidelines.^8^

A reviewer will independently extract the data in Covidence.^13^ A second trained reviewer will review the extracted data for accuracy. We will extract data on guideline details (first author or organisation, year, country), recommendations (screening, comprehensive assessment, treatment, other), the level of evidence supporting each recommendation, methods recommended to collect data and monitor falls prevention interventions, and consumer engagement.

### Guideline quality assessment

Two reviewers will assess independently the quality of the included clinical practice guidelines using both the Appraisal of Guidelines for Research and Evaluation Global Rating Scale (AGREE GRS) and the Appraisal of Guidelines of Research and Evaluation Recommendation EXcellence (AGREE-REX) tool.^14 15^ The two reviewers will meet to reach agreement about their rating scores. These two tools are recommended to be used together when completing a clinical practice guideline evaluation.^14^ The AGREE GRS tool has four key domains: (i) development process, (ii) style of presentation, (iii) comprehensiveness of reporting, and (iv) clinical validity.^15^ The AGREE-REX uses three factors to determine if a clinical practice guideline is high quality: (i) the credibility of the recommendations; (ii) involvement of relevant stakeholders and consumers in the development of the recommendations; and (iii) the degree to which the recommendations can be implemented.^14^

### Data analysis and synthesis

The data analysis and thematic synthesis will be completed independently by one reviewer in NVivo and discussed with the research team.^16^ The thematic synthesis will be divided into three main stages as instructed by Thomas and Harden ^17^. First, line-by-line coding will be performed on each included clinical practice guideline.^17^ Second, these codes will be compared across the clinical practice guidelines to develop descriptive themes that summarise a group of codes with similar meaning.^17^ Third, the descriptive themes will be interpreted to formulate analytical themes that transcend the content of the included guidelines to address any unanswered review questions.^17^

### Confidence in cumulative evidence

One trained reviewer will independently assess the overall strength of the review findings using the Grading of Recommendations Assessment, Development and Evaluation Confidence in Evidence from Reviews of Qualitative Research (GRADE-CERQual) framework to quantify the confidence reviewers can have in each review finding.^18^

## Supporting information

Example search strategy

## Data Availability

Data are available on reasonable request.

## ETHICS AND DISSEMINATION

Ethics approval is not required for the systematic review. The review will be submitted to an online peer-reviewed journal for publication.

### Contributors

JPM, CLP, A-MH, and MEM contributed to the design of the study and the formulation of the research questions. JPM wrote the draft of the protocol that was reviewed and edited by all authors. The protocol was approved by all authors prior to submission.

### Funding

This work was supported by an Australian Government Research Training Program Scholarship. And by the La Trobe University, Academic and Research Collaborative in Health.

### Competing interest statement

None declared.

### Patient consent for publication

Not required.

## Notes

### Competing Interest Statement

The authors have declared no competing interest.

