## Supplementary material for "Clinical practice guidelines to prevent hospital falls: protocol for a systematic review": Example search strategy

**Ovid MEDLINE(R) ALL**

| # | Searches |
|---|---|
| | **Hospital** |
| 1 | Hospitals/ (MeSH) |
| 2 | "Inpatient hospital*".mp. |
| 3 | "acute hospital*".mp. |
| 4 | hospitali?ation.mp. |
| 5 | Hospitalization/ (MeSH) |
| 6 | "acute care".mp. |
| 7 | Subacute Care/ (MeSH) |
| 8 | Inpatients/ (MeSH) |
| | **Falls** |
| 9 | 1 or 2 or 3 or 4 or 5 or 6 or 7 or 8 |
| 10 | Accidental Falls/ (MeSH) |
| 11 | fall*.mp. |
| 12 | "fall* prevention".mp. |
| 13 | "fall* management".mp. |
| 14 | "fall* risk*".mp. |
| 15 | (fall* adj15 injur*).mp. [mp=title, book title, abstract, original title, name of substance word, subject heading word, floating sub-heading word, keyword heading word, organism supplementary concept word, protocol supplementary concept word, rare disease supplementary concept word, unique identifier, synonyms, population supplementary concept word, anatomy supplementary concept word] (proximity operator) |
| 16 | (fall* adj15 fracture*).mp. [mp=title, book title, abstract, original title, name of substance word, subject heading word, floating sub-heading word, keyword heading word, organism supplementary concept word, protocol supplementary concept word, rare disease supplementary concept word, unique identifier, synonyms, population supplementary concept word, anatomy supplementary concept word] (proximity operator) |
| 17 | 10 or 11 or 12 or 13 or 14 or 15 or 16 |
| | **Clinical practice guidelines** |
| 18 | Practice Guideline/ (MeSH) |
| 19 | Practice Guidelines as Topic/ (MeSH) |
| 20 | "clinical practice guideline*".mp. |
| 21 | "clinical guideline*".mp. |
| 22 | Guideline Adherence/ (MeSH) |
| 23 | Evidence-Based Medicine/ (MeSH) |
| 24 | "best practice guideline*".mp. |
| 25 | "consensus statement".mp. |
| 26 | 18 or 19 or 20 or 21 or 22 or 23 or 24 or 25 |
| 27 | 9 and 17 and 26 |
| 28 | limit 27 to yr="1993 -Current" |
